# Prevalence of sleep disturbance and associated risk factors in UK Biobank participants with alcohol use disorders and major depression

**DOI:** 10.1101/2021.03.08.21252907

**Authors:** Bhanu Prakash Kolla, Joanna M. Biernacka, Meghna P. Mansukhani, Colin Colby, Brandon J. Coombes

## Abstract

**Introduction:** Current understanding of the differences in sleep disturbance (SD) and associated risk factors in patients with alcohol use disorders (AUD), major depressive disorders (MDD), and comorbid AUD+MDD is limited.

**Methods:** Data from the UK Biobank (UKB) (n=47,825) were utilized to categorize subjects into those with MDD (n=5,991), AUD (n=12,952), both (MDD+AUD)(n=3,219), and controls (n=25,663). We used generalized linear models (GLMs) to test whether rates of SD and sleep duration differed among the groups and determine the clinical predictors of SD. Rates of SD and sleep duration were compared using regression analyses accounting for demographic (age, sex, ethnicity, and Townsend deprivation index) and clinical (body mass index, neuroticism score, and alcohol consumption) factors.

**Results:** After accounting for diagnostic category, SD was associated with age, female sex, white ethnicity, and higher BMI, neuroticism and alcohol consumption scores (all p<0.0001).

The unadjusted prevalence of SD was 25.6%, 25.9%, 39.2%, and 41.1% in control, AUD, MDD, and MDD+AUD categories respectively. Rates of SD in controls and AUD group as well as MDD alone and MDD+AUD did not differ in unadjusted models (p=0.45 and 0.075, respectively). Prevalence of SD differed in the four groups (p<0.0001 for all pairwise comparisons) after adjusting for demographic confounders. After further adjustment for clinical factors, effect sizes were reduced, but pairwise comparisons remained significant, except in the AUD versus MDD group (all p<0.05). After adjusting for demographic and clinical factors, sleep duration did not differ among the groups.

**Conclusion:** Demographic and clinical characteristics associated with SD were similar in patients with MDD, AUD, and MDD+AUD. The differences in rates of SD between the diagnostic groups were attenuated but persisted after accounting for these confounders. Genetic and other factors capable of influencing SD in patients with MDD, AUD, and comorbid MDD+AUD merit future investigation.

## Introduction

Major depressive disorders (MDD) and alcohol use disorders (AUD) are among the most common psychiatric illnesses, with lifetime prevalence rates of 20.6% and 29% respectively (1, 2). These disorders are frequently comorbid; those with AUD have a 1.2-fold increased likelihood of having MDD in the previous year (2). The co-occurrence of these conditions is associated with increased severity of symptoms and overall worse prognosis (3, 4). Sleep disturbance (SD) is common in both conditions and when present, impacts associated morbidity, treatment response, and relapse risk (5-8). There is also evidence to suggest that SD is causally linked to alcohol dependence and is associated with certain circadian clock genes in those with AUD (9-12). However, the relative prevalence of SD in patients with AUD and/or MDD and its underlying etiological and associated risk factors are not well understood.

Prior studies of patients with AUD have shown that SD is associated with younger age, depressive symptoms, increasing severity and frequency of alcohol use, and psychosocial stress (7). In patients with MDD, the incidence of insomnia symptoms appears to increase with age (13, 14). SD occurring in the context of comorbid AUD and MDD has not been previously explored. The rates of SD in subjects with AUD, MDD, and both AUD+MDD in a single large population-based sample have not been compared before and research examining whether clinical and demographic factors associated with SD differ in each of these groups is lacking. Knowledge regarding the prevalence and factors associated with SD in patients with AUD and/or MDD could potentially help guide future screening, prevention, and treatment strategies for these conditions (15).

In this study, we aimed to utilize data from the UK Biobank (UKB) to examine the rates of SD and mean sleep duration in patients with AUD, MDD, and MDD+AUD. Further, we examined the association between SD and various clinical and demographic risk factors in each group. Finally, we investigated whether the prevalence of SD and self-reported sleep duration in these groups differed from controls after adjusting for associated demographic and clinical risk factors.

## Methods

### Population and setting

This study utilized data from the UKB, a large biomedical database with clinical and genetic information on approximately 500,000 individuals from across the United Kingdom, aged between 40 and 69 at recruitment (16).

### Sleep measurements and case definitions

Figure 1 summarizes the case and control definitions of SD, AUD, and MDD, as well as exclusion criteria.

**Figure 1:**
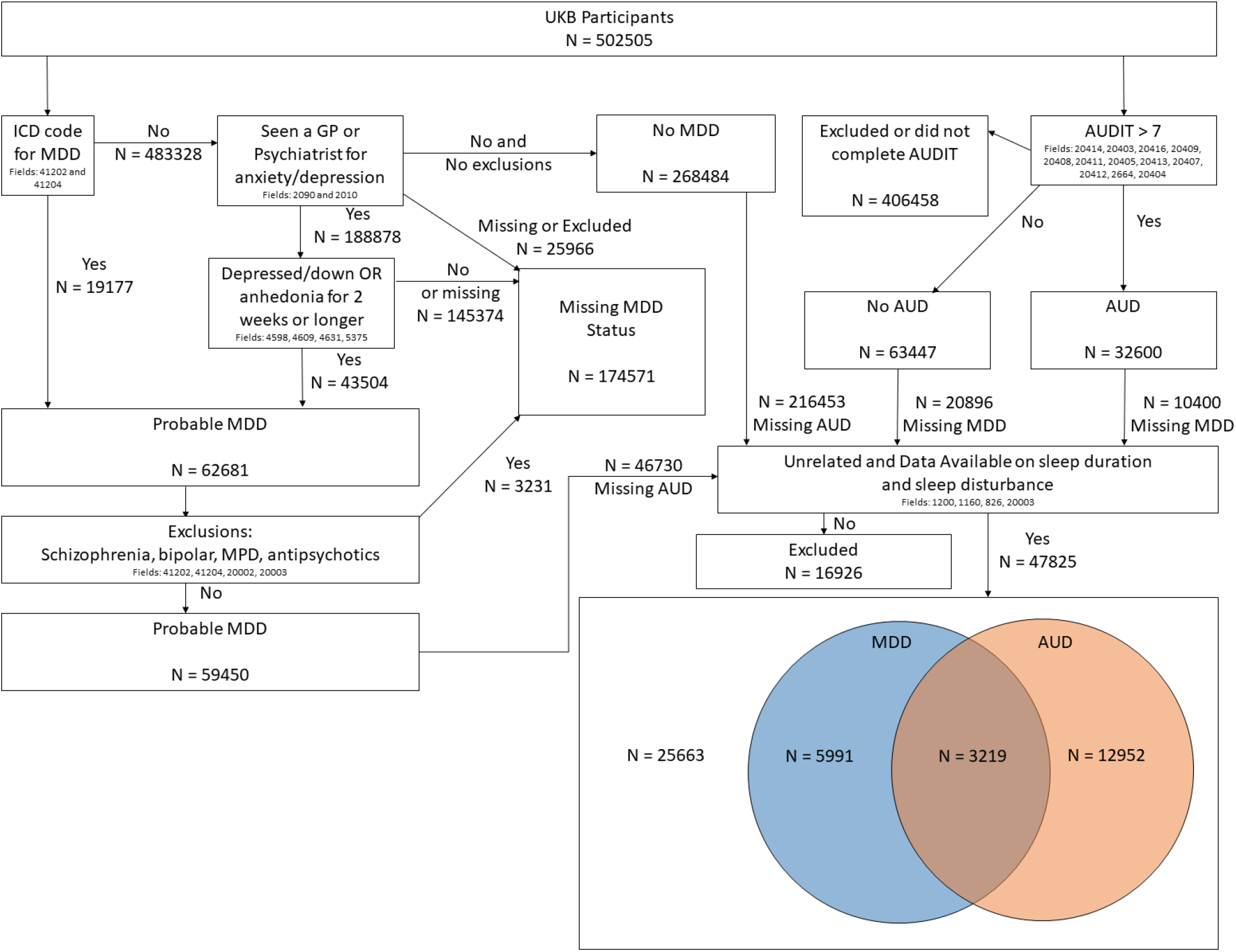
AUD/MDD Diagnosis group definitions derived from the UK Biobank Abbreviations: AUD = alcohol use disorders; AUDIT = Alcohol Use Disorders Identification Test; GP = general practitioner; ICD = International Classification of Disorders - 10^th^ Revision; MDD = major depressive disorders; MPD = multiple personality disorder; UKB = U.K. Biobank

SD was defined based on the response to the question: “Do you have trouble falling asleep at night or do you wake up in the middle of the night?” Those who responded “usually” were considered to have SD. Those who responded “never/rarely” or “sometimes” were considered to not have SD. Sleep duration was determined by subject self-report based on response to the question “About how many hours of sleep do you get in every 24 hours?”

Responses to the Alcohol Use Disorders Identification Test (AUDIT) were used to identify patients with AUD. An AUDIT score of ≥ 8 was considered to indicate AUD (17). About 100,000 participants completed the AUDIT, which was included an optional Mental Health Questionnaire (MHQ) (**Figure 1**). We defined participants as having “probable MDD” using criteria from previous studies, which also relied on completion of the MHQ (18)(**Figure 1**). Briefly, participants were classified as cases if they self-reported anhedonia or depression lasting longer than two weeks and had seen a general practitioner or a psychiatrist for anxiety/depression OR had a depression diagnosis (ICD-10 codes of F32, F33, F34, F38, F39, F40) in the medical record. The sample that completed the MHQ had similar prevalence rates of mental disorders when compared to a contemporaneous epidemiological study (19). To remove potential confounders of sleep with AUD and MDD, we excluded participants with a diagnosis of schizophrenia, bipolar disorder, personality disorder, and those currently taking antipsychotic medication(s). Finally, to ensure independence among the observations, we removed related individuals (second cousin or more related) by keeping only one participant from each family. Using these criteria, the cohort was divided into those with MDD alone, AUD alone, both (MDD+AUD), and controls with neither disorder (**Figure 1**).

### Additional data and measures

Demographic characteristics included age at enrollment (Field 21022), genetic sex (Field 22001), and self-reported ethnic group/ancestry (Field 22006). Townsend deprivation index (Field 189) was used as a proxy for socioeconomic status and was calculated immediately prior to the participant joining the UK Biobank utilizing the subject’s postal code. Positive values of the Townsend index indicate areas with high material deprivation, whereas negative values indicate relative affluence (20).

Clinical measures included a neuroticism score, weekly alcohol consumption, and body mass index (BMI). The neuroticism score was based on a subject’s response to 12 neurotic behavior domain questions and was a single integer score summing the number of “yes” responses to these 12 questions (Field 20127). The estimate of weekly alcohol consumption has been defined previously which calculates the average weekly alcohol intake by summing the self-reported consumption of white wine, spirits, beer, and fortified/red wine with a multiplied factor depending on the class of alcohol and removes extreme outliers (> 5 SD above the mean) (21). BMI was calculated from height and weight measured during the initial Assessment Centre visit (Field 21001).

### Analyses

We used generalized linear models (GLMs) to determine whether rates of sleep disruption differed among the groups. Specifically, we modeled SD or sleep duration as the outcome and an indicator for MDD/AUD group as the predictor. We tested whether sleep differed overall among the groups as well as whether there was evidence of pairwise differences between the groups. We considered models without adjustment for other covariates, adjusted for demographic factors (age at enrolment, sex, Townsend deprivation index, and race), and adjusted for demographic and clinical factors (BMI, neuroticism score, and alcohol consumption). Follow-up analyses were then performed to evaluate whether the effect of important demographic and clinical factors differed between the MDD/AUD groups, by testing the interactions effect between MDD/AUD groups and the covariates.

## Results

Requisite data were available for 47,825 subjects from the UKB, allowing them to be categorized into those with AUD, MDD, and controls. Based on our predetermined case definition criteria, the cohort was divided into those with MDD alone (n = 5,991), AUD alone (n =12,952), both (MDD+AUD) (n = 3,219), and controls (n = 25,663). Sample sizes varied slightly between analyses based on the amounts of missing data for clinical covariates.

### Clinical and demographic differences across diagnostic categories

Table 1 shows the unadjusted comparisons of demographic and clinical variables across the MDD/AUD groups.

**Table 1:** Distribution of sleep, demographic, and clinical variables among the four diagnostic groups.

| **Outcome** | **MDD**<br>(N = 5991)<br>Mean (SD) or N<br>(%) | **AUD**<br>(N = 12952)<br>Mean (SD) or N<br>(%) | **AUD+MDD**<br>(N = 3219)<br>Mean (SD) or N<br>(%) | **Controls**<br>(N = 25663)<br>Mean (SD) or N<br>(%) |
| --- | --- | --- | --- | --- |
| Insomnia | 2355 (39.2%) | 3360 (25.9%) | 1324 (41.1%) | 6572 (25.6%) |
| Sleep Duration | 7.18 (1.122) | 7.19 (0.916) | 7.15 (1.075) | 7.22 (0.941) |
| Age at Enrollment | 56.40 (7.392) | 54.63 (7.762) | 53.45 (7.445) | 57.87 (7.521) |
| Male | 1689 (25.1%) | 9868 (68.4%) | 1839 (49.8%) | 9622 (34.4%) |
| White | 5610 (83.4%) | 12606 (87.4%) | 3205 (86.8%) | 23048 (82.3%) |
| Townsend Deprivation Index (range -6.3 to 11) | -1.36 (2.89) | -1.62 (2.86) | -1.14 (2.99) | -1.87 (2.77) |
| Normal and Underweight (BMI < 25) | 2617 (39.0%) | 4539 (31.5%) | 1242 (33.7%) | 12343 (44.2%) |
| Overweight (BMI ≥ 25) | 2450 (36.5%) | 6905 (47.9%) | 1569 (42.6%) | 10696 (38.3%) |
| Obese (BMI ≥ 30) | 1637 (24.4%) | 2960 (20.5%) | 873 (23.7%) | 4903 (17.5%) |
| Neuroticism Score (range 0-12) | 5.79 (3.309) | 2.67 (2.449) | 6.16 (3.332) | 2.60 (2.401) |
| Alcohol Consumption (servings/week) | 7.75 (8.603) | 31.93 (18.176) | 29.55 (18.242) | 9.44 (8.152) |

Abbreviations: AUD = alcohol use disorders; BMI = body mass index, kg/m<sup>2</sup>; MDD = major depressive disorders; SD = standard deviation

Global tests of differences across the groups showed that the groups differed in age, sex, Townsend deprivation index, race, BMI, neuroticism, and alcohol consumption (all p < 0.0001). These differences were largely driven by differences of the control group compared to the three other case groups as seen in **Table 1**. Subjects in the control group were on average older, had lower mean Townsend index indicating less deprivation, a greater proportion of this group had normal BMI, and a smaller proportion were white individuals compared to the other 3 groups.

As expected, those with AUD (either in the comorbid MDD+AUD group or AUD alone) reported greater alcohol consumption, with 3-4 times greater consumption than those with MDD alone or controls. These groups also had a higher proportion of male, white, and overweight/obese individuals. Neuroticism scores of those with MDD (either in the comorbid MDD+AUD group or MDD alone) were on average twice as high as scores of those with AUD alone or controls.

### Sleep disturbance differences between diagnostic groups

The unadjusted prevalence of SD was lowest in the control group (25.6%) and highest in the MDD+AUD group (41.1%). The prevalence of SD in the MDD alone and AUD alone categories was 39.2% and 25.9% respectively.

Older age (p < 0.0001), white ethnicity (p = 0.009) and female sex (p < 0.0001) were associated with SD after adjusting for diagnostic category and other demographic factors.

The prevalence of SD in controls and the AUD alone group as well as the MDD alone and the MDD+AUD group did not differ in the unadjusted models (p=0.45 and 0.075, respectively). However, all four diagnostic groups had significantly different rates of SD (p < 0.0001 for all pairwise comparisons) after adjusting for demographic confounders (**Table 2**). The unadjusted rates of SD were found to be lower in the AUD groups (either AUD alone or MDD+AUD) as these groups had a much larger proportion of younger men.

**Table 2.** Effect estimates for predictors of sleep disturbance and comparisons of sleep disturbance between AUD/MDD groups in the UK Biobank. **Significant effects (p < 0**.**05) are denoted in bold**.

| Variable | Reference | Unadjusted<br>OR (95% CI) | Demographics<br>OR (95% CI) | Demographics +<br>Clinical<br>OR (95% CI) |
| --- | --- | --- | --- | --- |
| AUD | Controls | 1.019<br>(0.971,1.069) | **1.232 (1.169,1.297)** | **1.16 (1.077,1.249)** |
| MDD | Controls | **1.877 (1.769,1.99)** | **1.899 (1.788,2.015)** | **1.244 (1.144,1.352)** |
| MDD+AUD | Controls | **2.031 (1.883,2.19)** | **2.392 (2.212,2.586)** | **1.512 (1.363,1.678)** |
| MDD | AUD | **1.842**<br>**(1.726,1.965)** | **1.541 (1.44,1.65)** | 1.072 (0.972,1.183) |
| Both | AUD | **1.993 (1.839,2.16)** | **1.942 (1.79,2.107)** | **1.304 (1.183,1.436)** |
| Both | MDD | 1.082<br>(0.992,1.181) | **1.26 (1.153,1.377)** | **1.216 (1.084,1.363)** |
| Age | - | - | **1.024 (1.022,1.027)** | **1.029 (1.026,1.033)** |
| Male | Female | - | **0.715 (0.684,0.747)** | **0.719 (0.68,0.761)** |
| Townsend<br>Deprivation Index | - | - | 1.001 (0.994,1.009) | 0.997 (0.988,1.006) |
| White | Non-white | - | **1.08 (1.02,1.145)** | 1.048 (0.974,1.128) |
| BMI 25-29 | BMI < 25 | - | - | 1.023 (0.967,1.083) |
| BMI $\geq 30$ | BMI < 25 | - | - | **1.17 (1.09,1.255)** |
| Neuroticism Score | - | - | - | **1.136 (1.125,1.147)** |
| Alcohol<br>Consumption | - | - | - | **1.004 (1.002,1.005)** |

Abbreviations: AUD = alcohol use disorders; BMI = body mass index, kg/m<sup>2</sup>; MDD = major depressive disorders; OR = odds ratio; SD = standard deviation

Higher neuroticism score (p < 0.0001), alcohol consumption (p = 0.0002), and obesity (p < 0.0001) were associated with increased risk for SD after adjusting for diagnostic category and other demographic and clinical factors. Increasing age and female sex continued to be associated with SD after adjusting for diagnostic category and other demographic and clinical factors (both p < 0.05). Apart from the AUD alone versus MDD alone group comparison (p = 0.16), the pairwise comparisons remained significant (p < 0.0001) for SD after further adjusting for clinical risk factors, although with smaller effect sizes.

Finally, we tested whether demographic and clinical predictors of SD differed between the diagnostic groups by including an interaction between the predictor and the group indicator. Age and sex were the predictors of SD that significantly differed between diagnostic groups (interaction p-value = 0.007 and 0.0002, respectively). The age interaction was driven by a smaller effect size of age in the MDD alone group (OR = 1.01; p = 0.03) compared to all other groups (OR > 1.03; p < 0.0001 in all comparisons).

The sex interaction was driven by the fact that the effect of female risk of SD was smaller in the control group (OR = 1.21; p < 0.0001) than in any of the AUD/MDD diagnosis groups (OR > 1.4; p < 0.0001 in all comparisons). No other significant interactions were found.

### Sleep duration differences between diagnostic groups

The unadjusted self-reported sleep duration did not differ between the MDD, AUD and MDD+AUD diagnostic groups. The self-reported daily sleep duration in the control group was 7.22 ± 0.94 hours while the other groups reported 1.4 to 4.2 minutes of less sleep (**Table 3**). However, the differences in sleep duration between the controls and the other groups were not significant after adjusting for demographic and clinical factors.

**Table 3.** Effect estimates for predictors of sleep duration and comparisons of sleep duration between AUD/MDD groups in the UK Biobank. **Significant effects (p < 0**.**05) are denoted in bold**.

| Variable | Reference | Unadjusted<br>Beta (95% CI) | Demographics<br>Beta (95% CI) | Demographics +<br>Clinical Beta (95%<br>CI) |
| --- | --- | --- | --- | --- |
| AUD | Controls | **-0.03 (-0.06, -0.01)** | 0.01 (-0.02, 0.03) | -0.00 (-0.03, 0.03) |
| MDD | Controls | **-0.04 (-0.07, -0.01)** | -0.03 (-0.06, 0) | 0.03 (-0.01, 0.07) |
| MDD+AUD | Controls | **-0.07 (-0.10, -0.03)** | -0.03 (-0.06, 0.01) | 0.04 (-0.00, 0.09) |
| AUD | MDD | -0.01 (-0.04, 0.02) | **-0.03 (-0.06, -0.00)** | 0.03 (-0.01, 0.07) |
| AUD | Both | -0.04 (-0.07, 0.00) | -0.03 (-0.07, 0.01) | 0.05 (0.00, 0.09) |
| MDD | Both | -0.03 (-0.07, 0.01) | 0.00 (-0.04, 0.04) | 0.01 (-0.04, 0.06) |
| Age | - | - | 0.01 (0.00, 0.01) | 0.01 (0.00, 0.01) |
| Male | Female | - | **-0.06 (-0.08, -0.04)** | **-0.08 (-0.11, -0.06)** |
| Townsend<br>Deprivation Index | - | - | **-0.01 (-0.02, -0.01)** | **-0.01 (-0.01, -0.01)** |
| White | Non-white | - | **0.09 (0.06, 0.11)** | **0.08 (0.05, 0.11)** |
| BMI 25-29 | BMI < 25 | - | - | -0.02 (-0.04, 0.00) |
| BMI ≥ 30 | BMI < 25 | - | - | **-0.08 (-0.11, -0.05)** |
| Neuroticism score | - | - | - | **-0.02 (-0.03, -0.02)** |
| Alcohol<br>consumption | - | - | - | **0.001 (0.001, 0.002)** |

Abbreviations: AUD = alcohol use disorders; BMI = body mass index, kg/m<sup>2</sup>; MDD = major depressive disorders; SD = standard deviation

After adjusting for other clinical and demographic risks, male sex, non-white ethnicity, BMI ≥ 30, neuroticism score and lower alcohol consumption were associated with decreased sleep duration. The effect of age was moderated dependent on diagnosis group and age was not a significant predictor of sleep duration for those with MDD (either with or without AUD).

## Discussion

This is the first examination of sleep-related complaints in a single population-based sample comprising of subjects with AUD, MDD and MDD+AUD to our knowledge. The main findings were that the clinical and demographic factors associated with SD and sleep duration were similar in controls and across all diagnostic categories. After adjusting for diagnostic category, older age, female sex, white ethnicity, and greater BMI, neuroticism score and alcohol consumption were associated with SD. Patients with both AUD and MDD had the greatest sleep-related morbidity, with unadjusted and adjusted prevalence of SD significantly higher in this group compared to any other group. While the prevalence of SD in the AUD only category did not differ from controls in the unadjusted models, after adjusting for demographic and clinical factors, the prevalence was higher in the AUD group. Finally, after adjusting for both demographic and clinical factors, the prevalence of SD did not differ between the AUD only and MDD only groups.

In the general population, middle to older age has been shown to be associated with greater risk of insomnia symptoms, especially sleep maintenance difficulties (22), and women have been reported to have a 1.4-fold increased risk of SD compared to men (23). Lower socioeconomic status and psychological distress have also been found to be associated with SD (24),(25). On the other hand, in subjects with AUD, SD has been shown to be associated with a younger age, greater severity of AUD and alcohol use, and psychosocial stress (7). In our study, SD was associated with similar demographic and clinical factors across the different diagnostic categories, suggesting some degree of shared etiological and/or risk factors for SD in each of these conditions.

Further, when we examined whether the predictors of SD differed in terms of the magnitude of their influence in each of the groups, age showed a smaller effect in the MDD only group while sex showed a smaller effect in controls versus the other groups. The prevalence of insomnia symptoms was shown to increase with age in subjects with MDD in a single study where the baseline prevalence of insomnia was high at 77% in the 16-to-24 year age group and 90% in the 55-to-64 year age group (13). In our study, the prevalence of SD in the MDD group was 39%, higher than in controls and the AUD only group. The UKB included subjects were older, between the ages of 40-69 years at enrollment, which may have reduced the likelihood of detecting the effects of age on SD. Multiple previous studies have shown that the rates of SD are similar in men and women in early alcohol recovery (5, 6, 26, 27). The differential impact of sex on the prevalence of SD in AUD ± MDD compared to controls that we observed needs to be explored further in future studies.

Previous population-based estimates from the US and UK indicate that about 36% of adults endorse at least one insomnia symptom (13, 22). Data from our sample which utilized a single question demonstrated rates that were slightly lower (27.8%) in the controls, which may be partly due to the exclusion of those with MDD or AUD from this group. SD is highly prevalent in AUD in and the rates vary depending on whether patients are treatment seeking and the temporal relationship with last exposure to alcohol i.e. current active use, acute withdrawal, early recovery, or sustained recovery. The rates of SD in clinical samples are estimated to range between 36-91%, depending on the stage of AUD (5-7). SD is also extremely common in clinical samples with MDD, with a prevalence as high as 70-80% (14). In a population-based survey from the UK, 83.2% of subjects with MDD endorsed SD and 52% noted moderate severity of sleep-related symptoms (13). The overall prevalence of SD was lower in our sample compared to most of these previous studies, likely reflecting the different populations studied, especially in terms of age restrictions in the UKB, and different questions used to ascertain SD. To our knowledge, no previous studies have examined the prevalence of SD in subjects with comorbid AUD and MDD. Patients with MDD+AUD had greater rates of SD compared to controls and those with either condition alone in our study; this is in keeping with prior literature demonstrating that other morbidity is worse in patients with both conditions versus either condition (3, 4).

Neuroticism is a stable personality trait that has been shown to be associated with increased risk of both MDD and substance use disorders (SUD) and appears to predict MDD more strongly than SUD (28). Neuroticism has been linked with poor sleep quality and sleep habits (29-31). In our study, the neuroticism score was much higher in patients with MDD alone or MDD+AUD and was strongly predictive of SD after accounting for diagnostic category. Further, as expected, those with AUD reported much higher alcohol consumption than those with MDD alone and controls. Greater levels of alcohol consumption have also been shown to be associated with SD and short sleep duration (32, 33). The differences in the rates of SD between the diagnostic groups were attenuated as expected in our study after accounting for clinical factors, including neuroticism and alcohol consumption.

Our study has some limitations. The UKB sleep data relied on self-report and used a single question inquiring about sleep disturbance and duration. Our case definition for MDD utilized general practitioner diagnoses of ICD codes as well as self-reported depression/anhedonia, which could have led to misclassification of cases. Furthermore, our analysis of associated demographic and clinical risks was limited to a select set of measures that are known to affect sleep. After restricting our sample to patients who had the requisite data from the MHQ, we were left with 47,825 out of the total > 400,000 independent participants from the UKB, which may have biased the results. Recent investigations have revealed that those who completed the MHQ were better educated and of higher socioeconomic status; however, the rates of self-reported diagnoses have been found to be similar to population-based estimates (19). Even in light of these limitations, ours is the largest investigation of sleep disturbance and duration in a sample with AUD and MDD and first to show greater sleep morbidity in the AUD+MDD group.

## Conclusion

Clinical and demographic factors associated with sleep disturbance are similar across AUD, MDD, and MDD+AUD groups, suggesting potentially similar underlying risks and etiology. Those with MDD+AUD had worse sleep disturbance, further demonstrating that when these conditions co-occur they are associated with significantly greater morbidity. The lower prevalence of sleep disturbance in the AUD alone group prior to adjusting for demographic factors was likely influenced by overrepresentation of younger men in this category. Future studies should examine a more expansive range of demographic, clinical, and genetic factors to further understand the underpinnings of sleep disturbance in these disorders and identify potential etiological differences.

## Data Availability

This research has been conducted using the UK Biobank Resource under Application Number 55108.

## Acknowledgements

This research has been conducted using the UK Biobank Resource under Application Number 55108.

